# Cohort Profile: A longitudinal Victorian COVID-19 cohort (COVID PROFILE)

**DOI:** 10.1101/2023.04.27.23289157

**Authors:** Emily M. Eriksson, Anne Hart, Maureen Forde, Siavash Foroughi, Nicholas Kiernan-Walker, Ramin Mazhari, Erin C. Lucas, Mai Margetts, Anthony Farchione, Dylan Sheerin, George Ashdown, Rachel Evans, Catherine Chen, Shazia Ruybal-Pesántez, Eamon Conway, Marilou H. Barrios, Jasper Cornish, Maria Edmonds, Lee M. Henneken, Lisa J. Ioannidis, Sam W. Z. Olechnowicz, Ryan B. Munnings, Joanna R. Groom, Diana S. Hansen, Rory Bowden, Anna K. Coussens, Jason A. Tye-Din, Vanessa L. Bryant, Ivo Mueller

## Abstract

**Purpose:** The COVID PROFILE cohort is a longitudinal clinical study based in Victoria Australia, which was established to understand immunity to SARS-CoV-2 in a *low transmission population setting* and to identify immunological markers of long-term immunity and immune-dysregulation after both infection and vaccination. Additionally, this cohort was established as a biobank resource for researchers to address other health-related immunological questions.

**Participants:** We enrolled 178 adult community members, including household contacts, who had either recovered from a SARS-CoV-2 infection or were SARS-CoV-2 naïve. Only participants ≥18 years of age and, in the case of female participants, non-pregnant women at the time of enrollment were included in the study. Detailed COVID-19 clinical data, vaccination status, medical history and demographics was collected.

**Findings to date:** At enrollment, we found that 87.8% of COVID-19 recovered individuals were seropositive with detectable levels of anti-SARS-CoV-2 IgG antibodies. Seronegative COVID-19 recovered individuals included asymptomatic individuals or participants that were enrolled more than 12 months after their COVID-19 diagnosis. Except for one individual who was seropositive at baseline despite a previous SARS-CoV-2 PCR negative diagnosis, all household contacts and other community members enrolled as SARS-CoV-2 PCR negative, were seronegative for all SARS-CoV-2-specific antibodies tested. The infection rate (re-infection or new infection) during 24 months of the study was 42.7%, as determined by either rapid antigen tests, PCRs or serology screens. Of the SARS-CoV-2 recovered participants, 32.6% reported ongoing symptoms at enrollment of which 47% had already experienced ongoing symptoms for more than 12 weeks.

**Future Plans:** COVID PROFILE will be used to comprehensively understand temporal immunity to SARS-CoV-2 and COVID-19 vaccines and to understand the impact of host immunological composition on such immunity and symptom severity. Additionally, studies focusing on understanding immunity following breakthrough infections and immunological risk factors that contribute towards development of long COVID are planned.

**Limitations/Strengths of the study:**

- Extensive clinical information is available and longitudinal samples (blood, saliva and oropharyngeal swabs) collected at regular intervals up to 2.5 years after initial enrolment.
- This low SARS-CoV-2 transmission population cohort, enables exploration of difference in both the acquisition and maintenance of naturally acquired and vaccine-induced immunity not confounded by prior, frequent and/or undetected exposures.
- Extensive biobank of numerous blood fractions and biospecimen enables further exploration of mucosal immunity, nasal microbiome and humoral and cellular immunity over time.
- The cohort may be limited in addressing research questions regarding outcomes and risk factors and their associations where low incidence is expected.

## Introduction and Cohort description

The first case of SARS-CoV-2 recorded in Australia was on January 25^th^ 2020, only 25 days after the first case had been reported to the World Health Organisation (1). This was followed by intense national efforts to diminish and if possible, eliminate SARS-CoV-2 transmission. While transmission was not immediately contained and resulted in a peak of confirmed SARS-CoV-2 positive cases in late March followed by a second peak in July, community transmission was largely undetectable by October 2020 (2) (**Fig. 1**). This pandemic response, which included international and domestic border closures, lockdowns and Australia’s geographical inaccessibility resulted in overall comparatively low case numbers and thus a distinctively SARS-CoV-2 naïve population prior to COVID-19 vaccine rollout in February 2021. Furthermore, unlike the majority of populations across the world, the Australian population remained largely unexposed until end of 2021 (**Fig.1**).

**Figure 1.**
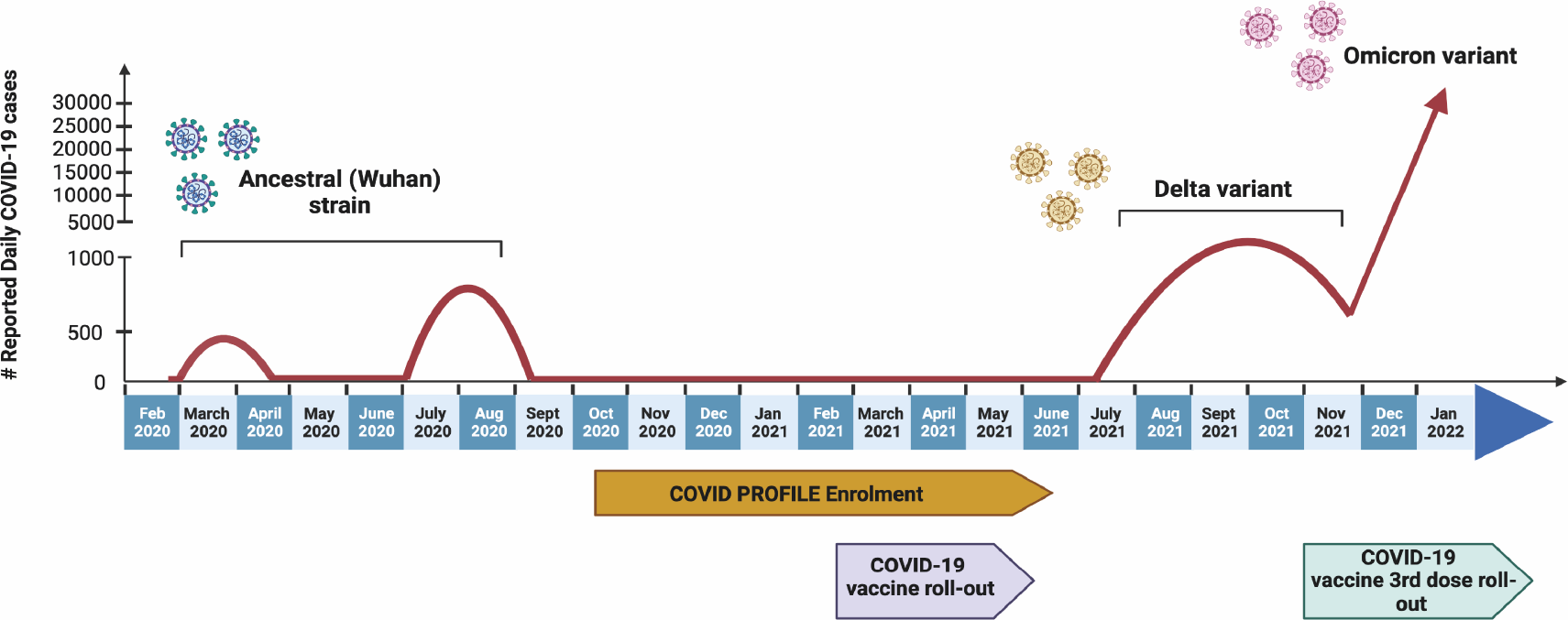
Schematic of COVID-19 pandemic in Australia 2020-2022. Schematic timeline showing daily reported cases in Australia (2, 10) between Feb 2020 to Jan 2022 with indicated dominant infective variant identified in reported cases over time (9). Created with BioRender.com

The COVID PROFILE study was launched in Melbourne, Victoria in October 2020 to follow COVID-19 recovered individuals over a year of enrollment, being in a relatively unique position to understand longevity of immunity to SARS-CoV-2 infection where risk of continuous re-exposure was low. This study was approved by the Walter and Eliza Hall Institute (WEHI) and Melbourne Health Human Research Committees (Projects 20/08 and RMH69108 respectively). All enrolled participants provided written informed consent to participate in this study, in accordance with the Declaration of Helsinki. Most participants were enrolled between October 2020 and June 2021. SARS-CoV-2 recovered participants were predominantly infected locally during the second July-September 2020 infection wave (71.4%) and the remaining participants contracted SARS-CoV-2 from the first infection wave (March-May 2020) or were infected overseas (**Fig. 1**). The only circulating SARS-CoV-2 variant at the time of these infections was the ancestral (Wuhan) strain resulting in a homogenously exposed population (**Fig. 1**). The study visit schedule consisted of a baseline visit (at enrollment), 4-6 weeks post-diagnosis where possible, followed by visits at 3, 6, 9 and 12 months post-enrollment (**Fig. 2**). Following the roll out of COVID-19 vaccines in Australia in Feb 2021, that consisted predominantly of ChAdOx1-S (AstraZeneca) or BNT162b2 (Pfizer), participants already enrolled in the study were invited to continue follow-up for up to 12 months after their second dose of vaccine. Additional SARS-CoV-2 naïve individuals were invited to take part in the study before their first COVID-19 vaccination and for these newly recruited participants, a baseline pre-vaccination sample was also collected. Post-vaccination follow-up visits consisted of sample collection 2-4 weeks after each dose of vaccine and then 3, 6, 9 and 12 months after the second dose, unless another exposure event such as an additional vaccination or breakthrough infection occurred. Follow-up was then extended for up to 12 months after each exposure, following a visit schedule comparable to that previously defined (**Fig. 2**). Biospecimens consisting of blood, saliva, and oropharyngeal swabs were collected at all visits where blood specimens were stored as serum, plasma, cryopreserved peripheral blood mononuclear cells (PBMCs), frozen and fixed whole blood and in some cases neutrophils. Oropharyngeal swabs were collected and stored in sample preservation medium in their original tubes at -80°C until required. Detailed COVID-19 clinical data, vaccination status, medical history and demographics (both during acute infection and long-term follow up) was collected and study data were managed using REDCap electronic data capture tools hosted by the Clinical Translation Centre, WEHI, Parkville, Victoria, Australia (3, 4).

**Figure 2.**
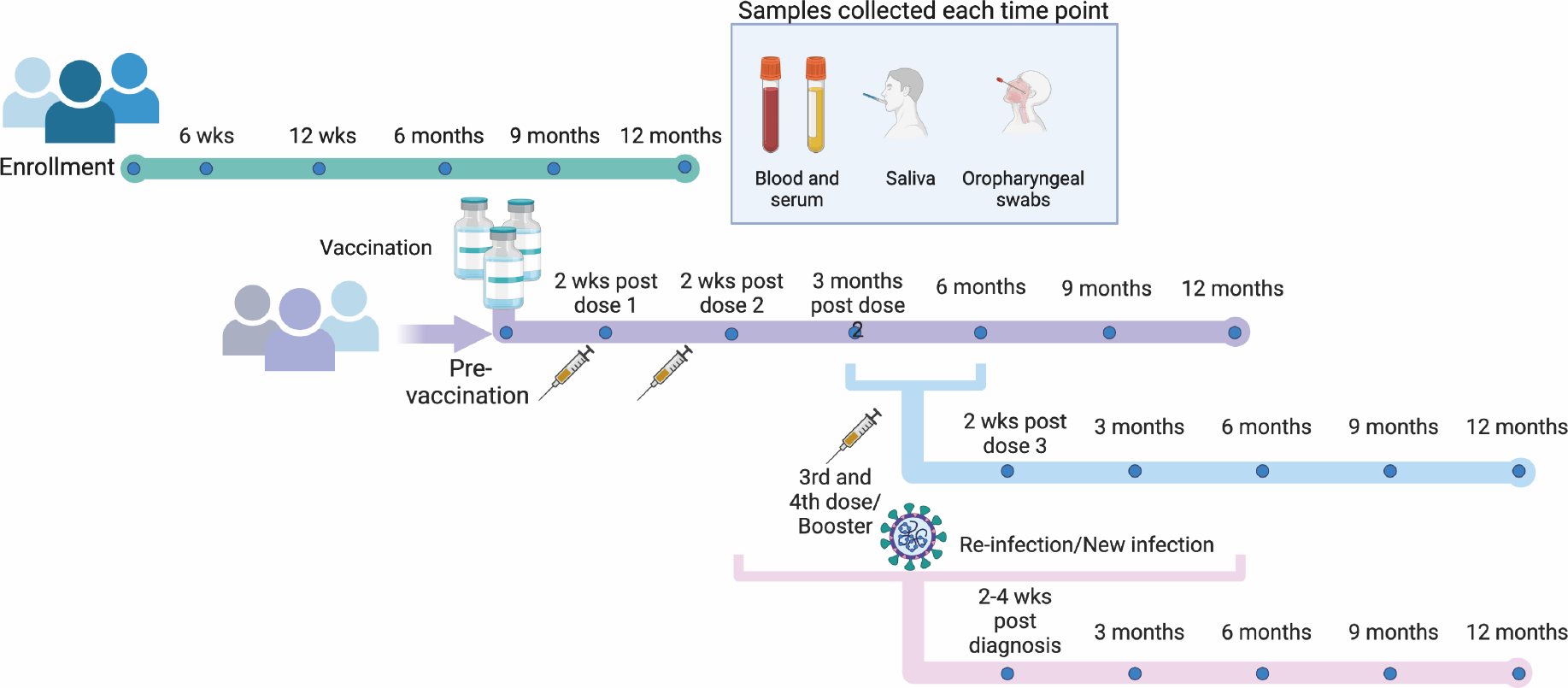
Study design and participant visit schedule. SARS-CoV-2 recovered participants were enrolled and followed for up to 12 months after enrollment with sample collection at baseline (at enrollment) and 3, 6, 9 and 12 months after enrollment (green line). SARS-CoV-2 naïve participants provided a baseline sample (purple line). Biospecimen samples were collected from COVID-19 vaccinated participants 2-4 weeks after each vaccine dose and then at 3, 6, 9 and 12 months after the final dose (purple and blue line). Samples were also collected 2-4 weeks after breakthrough infections and again at 3, 6, 9 and 12 months after the diagnosis (pink line). Saliva, oropharyngeal swabs, serum and whole blood were collected and stored after each visit. Created with BioRender.com

### Participant demographics

To date, 178 participants have been recruited to the COVID PROFILE study following advertising campaigns via social media, primary care networks and tertiary teaching hospital. Seven participants failed to provide a baseline sample as specified in the study schedule and were excluded from the study and all analyses (**Fig. 3**). Ninety-eight participants were SARS-CoV-2 recovered and 73 were SARS-CoV-2 naïve at enrolment. The participants comprised 122 separate households: 88 households with 1 participant, 26 households with 2 participants, 1 household with 3 participants and 7 households with 4 participants. Gender distribution was 59.1% female (n = 101) in the overall cohort and 63.3% female (n = 62) in the COVID-19-recovered group. Overall age distribution in the cohort ranged from 18 to 90 years of age with a median age of 46 years. Median age for the SARS-CoV-2 naïve group was 44 years (range 18 – 90 years) and 49 years for the SARS-CoV-2 recovered group (range 19 – 75 years, **Fig. 4)**, which resemble the overall distribution of nationwide adult cases in 2020 (5).

**Figure 3.**
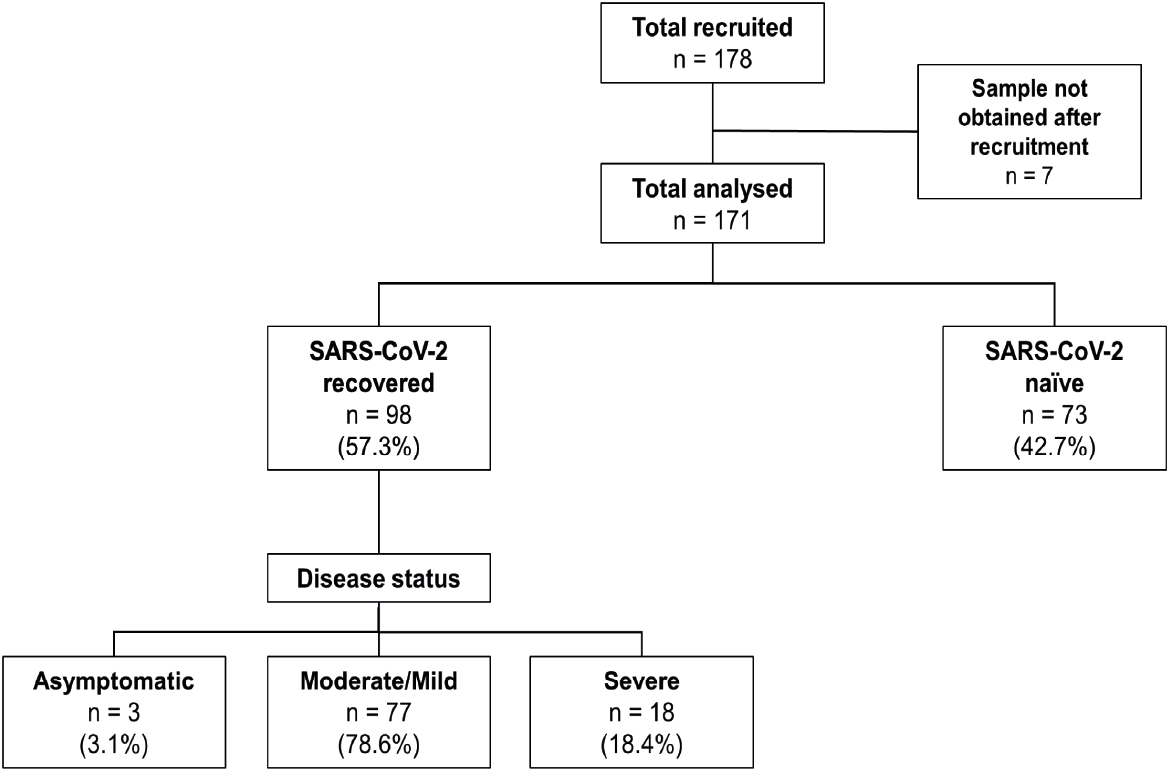
Summary of participant categories. Cohort participants stratified based on infection status at enrollment. Further stratification of SARS-CoV-2 recovered individuals based on disease status.

**Figure 4.**
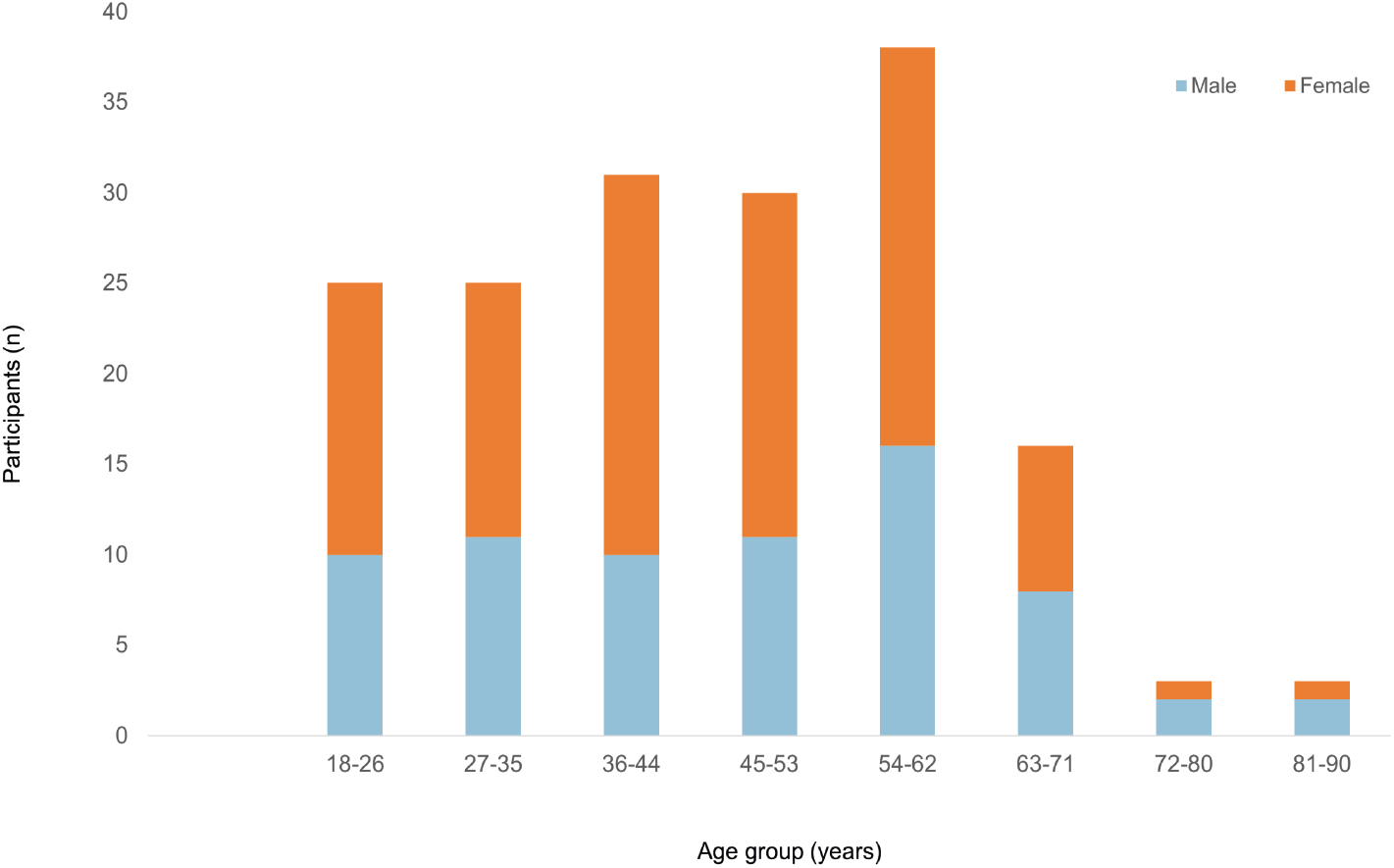
Cohort age and gender profile. Participant age range is 18-90 years; blue: male; orange: female.

Self-reported acute disease symptoms and duration were collected at enrolment and after subsequent breakthrough infections. Acute disease status was classified as “severe disease” if hospitalisation was required due to the infection, regardless of whether intubation or oxygen supplementation was necessary. All other participants with any symptoms were categorised as mild/moderate (78.6%; n=77) or asymptomatic (3.1%; n=3) in cases of self-reported absence of symptoms (**Fig. 3**). A substantial proportion of participants were categorised as severe (18.4%; n=18), which was higher than the reported national rate of 12.5% hospitalisation in 2020 due to COVID-19 (6).

#### COVID-19 vaccine status

During follow up most participants received at least 2 doses of COVID-19 vaccines (81.9%). The breakdown of vaccine type received as primary vaccination is summarised in **Fig. 5**. Participants predominately received Pfizer (60%, median age = 40.5 years) and AstraZeneca (37.1%, median age = 56.5 years) with only a few receiving Moderna-Spikevax (0.7%) or Novavax (2.1%). A subset of the vaccinated participants have also received a third (43.4%) and fourth (35.7%) dose of vaccine of either Pfizer or Moderna-Spikevax.

**Figure 5.**
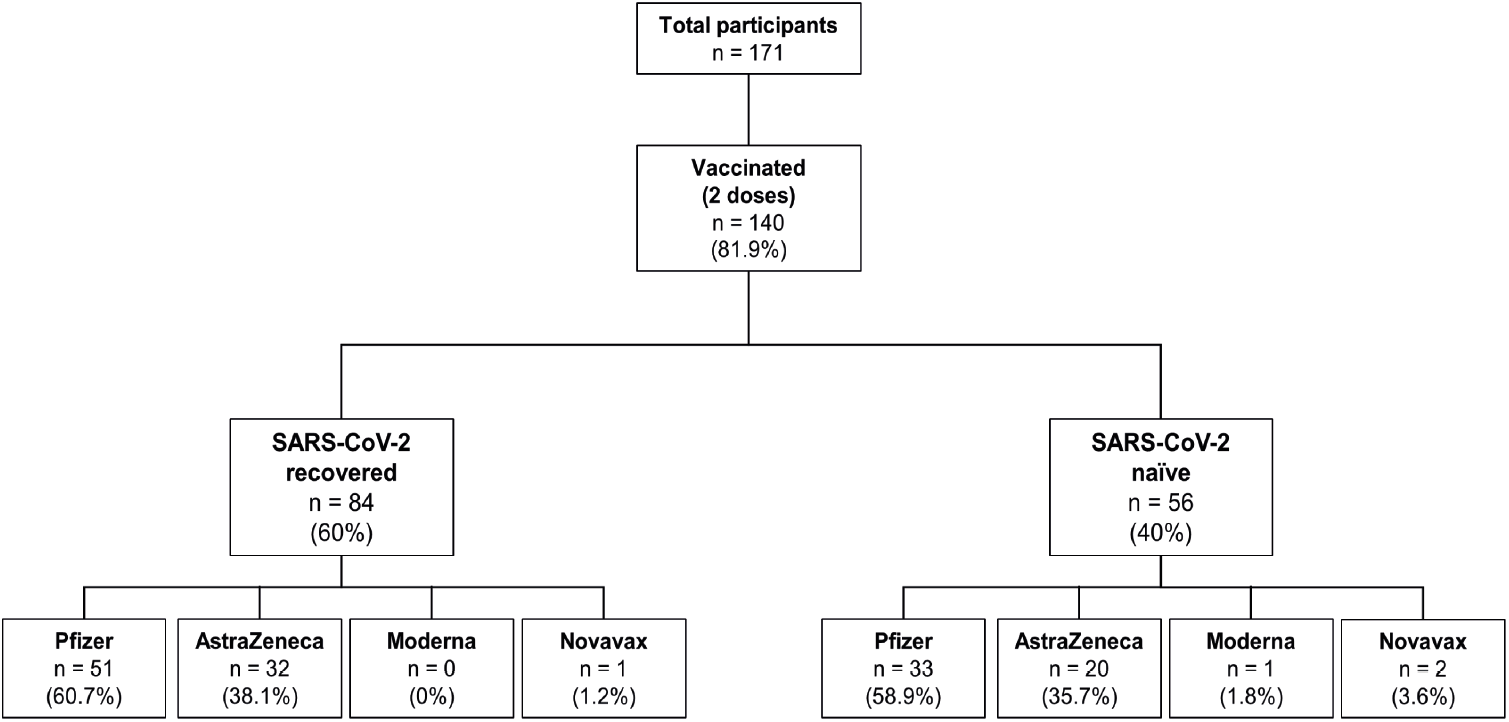
Summary of vaccine status of participants. Cohort participants stratified based on disease status at time of enrollment and vaccine status after two doses of COVID-19 vaccine. Vaccine recipients were further stratified based on the type of COVID-19 vaccine received; Pfizer (BNT162b2), AstraZeneca (ChAdOx1-S), Moderna-Spikevax or Novavax.

### Findings to date

To establish the serostatus of participants, SARS-CoV-2-specific IgG antibodies were measured at baseline for all participants using a Luminex multiplex assay (7). This assay measures antibodies that recognize and bind to full-length Spike, receptor-binding domain (RBD), S1, S2 and nucleoprotein, all based on the Wuhan variant sequence. In addition, antibodies specific for seasonal coronavirus proteins (NL63, HKU1, OC43 and 229E), tetanus toxoid and Influenza A (H1N1) were also measured as controls. Serostatus based on measured anti-SARS-CoV-2 antibody levels was determined using a random forest classification algorithm. Most participants who had a PCR+ diagnosis at enrolment were seropositive (87.8%) at baseline. Amongst the COVID-19 recovered individuals who had a PCR+ diagnosis but were seronegative at enrolment (n=12), three presented as asymptomatic individuals, four participants were enrolled more than 12 months after their COVID-19 diagnosis and the remaining seronegative individuals were within 6 months of diagnosis. The majority of individuals that were enrolled as PCR negative participants (99%) were also seronegative, with only one participant who had previously had a negative PCR diagnosis shortly before sample collection, was found to be seropositive.

As part of the extensive longitudinal clinical data collected for this study, a substantial proportion of SARS-CoV-2 recovered participants (32.6% of 98 SARS-CoV-2 recovered individuals) reported ongoing symptoms at the time of enrolment of which 47% had already experienced symptoms persisting for 12 weeks or longer, meeting the current criteria for long COVID (8).

The longitudinal design of our study captured multiple breakthrough infections (42.4%) in our participants over the course of 24 months. Most such detections came from rapid antigen tests (RATs), precluding the identification of the infecting variant. However, 98.3% of breakthrough infections coincided with the period during which Omicron was the dominant variant in Australia (9). Most breakthrough infections were detected after three COVID-19 vaccine doses (65.5%) but they also occurred after two (15.5%) and four (19%) doses of COVID-19 vaccines

### Future plans

Sample collection is expected to be completed in June 2023 after which samples will support in-depth analysis to understand temporal humoral and cellular responses to infection, COVID-19 vaccines and breakthrough infections with different variants. We have so far collected and bio banked > 8300 biospecimens, which include serum, whole blood, oropharyngeal swabs, plasma, PBMCs and saliva samples across 1200 visits over 24 months. Furthermore, given the detailed medical history recorded and the relatively high frequency of participants reporting long COVID in our cohort, additional analyses are planned to explore specific biomarkers and underlying processes involved with this condition.

### Patient and public involvement

The participants were not involved in the study’s design or conclusions. Participants were informed of serological results via email.

### Limitations/Strengths

The COVID PROFILE is a cohort study where extensive clinical information is available and longitudinal blood samples, saliva and oropharyngeal swabs collected at regular intervals up to 2.5 years after initial enrolment will have been accumulated at the completion of the study. This included samples collected in naive individuals with subsequent infection. Low SARS-CoV-2 transmission in Australia before January 2022, provides an opportunity to explore difference in both the acquisition and maintenance of naturally acquired and vaccine-induced immunity (or a combination thereof), as well as differences in vaccine response in people with a single or no previous exposure to SARS-CoV-2. This also provides a unique opportunity to support research questions not confounded by prior, frequent and/or undetected exposures using a highly curated and clinically annotated biobank of samples. We anticipate the COVID PROFILE cohort will also provide great value as a comparative cohort set to other studies in high transmission countries. Our extensive biobank includes numerous blood fractions and biospecimen types collected longitudinally, which further enables the exploration of mucosal immunity, nasal microbiome and humoral and cellular immunity over time. However, due to its relatively small size, the cohort may be limited in addressing research questions regarding outcomes and risk factors and their associations where low incidence is expected.

#### Collaboration

The authors are welcoming and encouraging research collaborations using the COVID PROFILE samples for population and/or immunological questions. Samples and/or data are available on reasonable request and researchers are welcome to contact the research group for further information.

## Data Availability

All data produced in the present study are available upon reasonable request to the authors

## Data availability statement

Samples and/or data are available upon reasonable request. Research proposals for future collaboration are welcome. Applicants for collaboration and more information are encouraged to contact the corresponding author Dr. Emily M Eriksson (Email address).

## Author Contributions

EME and IM conceived and initiated the study. EME, IM, VLB and JATD were the principal investigators of the study. EME established study protocols and oversaw clinical, technical and data analysis staff. EME, RB and AKC established sample processing procedures. EME and SF wrote the manuscript. AH and MF were study coordinators managing recruitment, enrolment, collected samples and obtained clinical and demographic data. SF managed the Redcap database. NKW, RM, ECL, MM, AF, DS, GA, RE, CC, MHB, JC, LJI, SWZO, and RBM provided technical assistance. ME and LMH assisted with sample collection. SF, SRP and EC managed data analysis. IM, VLB, JATD, JRG, DSH, RB and AKC critically reviewed the manuscript. All authors read and approved submission of the manuscript.

## Acknowledgments

We gratefully acknowledge all the participants of this project and St Vincent’s hospital nurses for assistance with home visits and sample collection. We also thank all administrative and communications staff and personnel for their support.

